# The emotional impact of gambling-related advertising: an experimental functional Near-Infrared Spectroscopy study protocol

**DOI:** 10.64898/2026.05.20.26353682

**Authors:** Abraham Ros-León, Sergio Molina-Rodríguez, Olga Pellicer-Porcar, Victor Cabrera-Perona, Joaquín Ibañez-Ballesteros, Daniel Lloret-Irles

**Affiliations:** Department of Health Psychology, Miguel Hernandez University; Department of Physiology, Miguel Hernandez University

**Author notes:** Corresponding author: Daniel Lloret-Irles.

**Keywords:** Advertising, Gambling, fNIRS, Neuroimage, Study Protocol

## Abstract

**Background:** The proliferation of gambling advertising has intensified concerns regarding its influence on vulnerable populations, yet the neural mechanisms underlying cue-reactivity to these stimuli remain underexplored in ecologically valid settings. This study protocol proposes a novel methodological framework to investigate prefrontal cortical responses to gambling advertisements in individuals with varying degrees of gambling experience.

**Materials and methods:** This cross-sectional study will recruit 44 participants, divided into a clinical group (individuals with high-frequency gambling or gambling disorder) and a matched control group. Neural activity will be recorded using fNIRS while participants view gambling-related, neutral, violent, and sexual stimuli. Secondary measures include validated scales for gambling severity (SOGS), impulsivity, sensation seeking, and alexithymia. Data analysis will primarily utilize inter-subject correlation (ISC) to quantify neural synchronization and multiband frequency decomposition to capture dynamic affective processing. Advanced preprocessing, including short-channel regression, will be applied to ensure signal robustness.

**Discussion:** By combining portable neuroimaging with a data-driven ISC approach, this study aims to identify objective neural markers of gambling vulnerability. The findings will provide novel insights into the idiosyncratic processing of commercial stimuli, potentially informing public health policies and the development of more effective evidence-based regulations for gambling marketing.

## Background

In recent years, the gambling industry has experienced a notable expansion that has been accompanied by substantial increase in advertising and marketing strategies, raising growing concerns about their potential impact on public health. Gambling advertisements are designed to capture attention, enhance emotional engagement, and promote participation, often reaching individuals who may be particularly vulnerable to developing problematic gambling behaviors (^1, 2, 3^). Understanding how these commercial stimuli influence cognitive and affective processes is therefore critical for assessing their role in the development and maintenance of gambling-related problems.

Advertising plays an important role in stimulating participation in gambling, both among the general population (^4^), as well as among young people (^5, 6^) and problem gamblers (^7^), and thus increasing the likelihood of developing gambling disorder. The meta-analysis by Bouguettaya et al. (^8^) suggests a positive association between exposure to gambling advertising and attitudes toward betting, as well as gambling intentions and behavior. Some of the key mechanisms through which gambling advertising operates include fostering a positive attitude toward gambling and a favorable social perception (^9; 10; 11^).

Since the publication of the DSM-V in 2013, the American Psychiatric Association has recognized gambling disorder as a behavioral addiction. According to the latest version (^12^), pathological gambling –recognized as gambling disorder– is a behavioral condition characterized by persistent loss of control over gambling activities, despite significant negative personal, social, and financial consequences, and it is classified among addictive disorders due to its clinical similarities. In terms of prevalence among adults, recent meta-analysis suggests a prevalence in Western Europe of approximately 17% (ranging from 9.9 to 25.6%) for at any risk gambling, and around 2.6% (ranging from 1.8 to 3.4%) for problem gambling (^13^). At the Spanish level, it is estimated that 1.4% of adults present one or more criteria of the DSM-5 for gambling disorder (^14^).

Gambling-related cues such as images, sounds, or narratives commonly used in advertisements, can trigger conditional responses, including craving, attentional bias, and increased motivation to gamble. These processes, widely documented in substance-related addictions, have also been observed in gambling disorders and provide a useful framework for understanding how advertising stimuli may impact cognitive and affective processing.

In line with this framework, there is a limited body of research employing neuroimaging techniques to investigate the neural and cognitive effects of gambling advertising. Among the available studies, there is a marked predominance of functional magnetic resonance imaging (fMRI). Although the overall number of such studies remains relatively small, existing evidence points to the involvement of frontal brain regions in cue-reactivity processes. In particular, the study by Crockford et al. (^15^) reported increased activation in the dorsolateral prefrontal cortex during exposure to gambling-related stimuli, which has been interpreted in terms of cognitive control and reward evaluation mechanisms. Similarly, studies such as those by Potenza et al. (^16^) and Goudriaan et al. (^17^), although not exclusively focused on frontal areas, further support the involvement of fronto-cingulate networks, especially in relation to craving and the motivational salience of cues.

Despite the findings mentioned, fMRI-based research presents important limitations regarding temporal resolution and ecological validity. The BOLD signal measured by fMRI reflects hemodynamic changes that unfold over several seconds, which limits the ability to capture faster neural dynamics and rapid cognitive processes involved in decision-making and affective responses (^18^). In addition, the ecological validity of fMRI studies is often constrained by the highly controlled and artificial scanning environment, where participants must remain motionless inside a noisy and confined space. Such conditions can affect natural behavior, emotional processing, and attention (^19, 20, 21, 22^). In this context, near-infrared spectroscopy (fNIRS) emerges as a promising methodological alternative.

fNIRS is grounded in the premise that the activation of a brain region leads to an increase in local oxygen consumption and, as a regulatory response, a subsequent increase in blood flow to that region. This phenomenon, known as “neurovascular coupling,” results in an increase in oxyhemoglobin (HbO) concentration and a decrease in deoxyhemoglobin (HbR). These changes in hemoglobin concentration are detected by the NIRS instrument as variations in the absorption of the infrared light it emits (^23, 24^). Compared to fMRI, fNIRS provides improved temporal sensitivity, allowing a more precise characterization of the dynamic processes underlying cognitive and affective responses. Moreover, its portability and relative tolerance to motion enable data acquisition in more naturalistic settings, thereby enhancing ecological validity (^25, 26^). These features are particularly relevant in the study of gambling behavior, where decision-making and cue-reactivity processes typically unfold in dynamic, real-world environments that are difficult to replicate within the constraints of an fMRI scanner. However, capturing neural responses in such dynamic and naturalistic contexts also requires analytical approaches capable of identifying shared and stimulus-driven patterns of brain activity across individuals.

Inter-subject correlation (ISC) is a data-driven approach that quantifies the synchronization of brain activity across individuals exposed to the same stimuli, without requiring a priori assumptions about the temporal structure of neural responses (^27, 28^). Originally developed in fMRI research, ISC has been widely used to study neural responses to complex and naturalistic stimuli, demonstrating that shared inputs can elicit temporally aligned activity across participants (^27, 29^). Importantly, ISC is sensitive to variability in higher-order cognitive and affective processing: while sensory regions typically show high synchronization, prefrontal areas often exhibit lower consistency, reflecting more individualized responses (^30, 31^). Furthermore, Finn et al. (^32^) have shown that ISC is not only driven by shared stimulus properties, but is also sensitive to certain individual psychological characteristics. Specifically, their recent work has demonstrated that trait paranoia modulates ISC, with individuals exhibiting more heterogeneous ISC within trait-defined groups than between groups. This feature makes ISC particularly suitable for investigating responses to emotionally salient stimuli, as different emotional contents can modulate the degree of inter-subject synchronization and engage prefrontal regions involved in affective processing (^33, 34^).

Finally, while previous research has demonstrated the utility of fNIRS to examine the capacity of advertising stimuli to engage audiences (^35^) and to decode affective states more broadly (^36^), the combination of fNIRS with ISC analysis remains unexplored in the context of gambling-related advertising. To the best of our knowledge, and based on current meta-analytic evidence, this study is the first to employ this methodology to investigate emotional responses within this specific domain. This gap in the literature highlights the need for methodological approaches capable of capturing both the dynamic and ecological nature of such stimuli. In this context, the present study proposes a novel protocol combining fNIRS and advanced signal analysis techniques to examine prefrontal responses to gambling advertisements in individuals with and without gambling-related problems. Therefore, the primary aim of this study is to investigate the neural and behavioral responses to gambling-related advertising using fNIRS combined with ISC analysis. Specifically, the objectives and hypothesis are as follows:

O1: To examine differences in ISC between individuals with high frequency gambling behavior or gambling disorder and non- or low-frequency gamblers during exposure to gambling-related stimuli.

H1: Individuals with high frequency gambling behavior or gambling disorder will exhibit significantly lower ISC values than non- or low-frequency gamblers during exposure to gambling-related stimuli.

O2: To examine differences in ISC between individuals with high frequency gambling behavior or gambling disorder and non- or low-frequency gamblers during exposure to neutral, violent and sexual stimuli.

H2: No significant differences in ISC will be observed between individuals with high frequency gambling behavior or gambling disorder and non- or low-frequency gamblers during exposure to neutral, violent, and sexual stimuli.

O3: To assess the predictive capacity of psychological variables associated with gambling behavior on ISC during exposure to gambling-related stimuli.

H3: Psychological variables associated with gambling behavior will significantly predict the ISC values during exposure to gambling-related stimuli.

O4: To evaluate the within-group variability of ISC in individuals with high frequency gambling behavior or gambling disorder and non-gamblers or low frequency gamblers.

H4: ISC variability will be significantly greater in individuals with high frequency gambling behavior or gambling disorder than in non- or low-frequency gamblers.

## MATERIAL AND METHODS

### Study design and setting

This study will employ a cross-sectional comparative design combining psychological and neurobiological measures. Psychological assessment will be conducted using validated self-report questionnaires and scales, while neurobiological data will be obtained through functional near-infrared spectroscopy (fNIRS).

This study protocol has been developed in accordance with current methodological recommendations for fNIRS research, as outlined by the Society of Functional Near-Infrared Spectroscopy (^37^). All procedures related to signal acquisition, preprocessing, and reporting will adhere to these guidelines to promote transparency, reproducibility, and comprehensive methodological reporting.

### Ethical considerations

This study protocol was approved by the Ethics Committee for Research Involving Medicines at the University Hospital of San Juan - Alicante (Cod. 19/334 Tut.) and the Research Ethics Integrity Committee of Miguel Hernández University (DPS.DLI.241115). This study will be conducted in accordance with the Declaration of Helsinki, and all data will be managed in compliance with the Organic Law 3/2018 on the Protection of Personal Data and Guarantee of Digital Rights. Participation is voluntary, and participants will provide written informed consent before being enrolled. Data confidentiality will be maintained throughout the research process.

### Sample size estimation

This strategy is consistent with the study’s objective and exploratory nature, as well as with the practical limitations of clinical recruitment.

We did not rely on an a priori statistical power analysis, as such approaches may overestimate required sample sizes due to publication bias (^38, 39^), as well as there is a limited body of research on fNIRS and emotional reactivity in pathological gamblers exposed to emotional and gambling-related stimuli. Instead, we conducted a sensitivity power analysis to determine the minimum effect size detectable assuming a statistical power of 0.80 and an alpha level of 0.05. This strategy is consistent with the study’s objective and exploratory nature, as well as with the practical limitations of clinical recruitment. Using G*Power 3.1.9.6. (^40^), the analysis indicated that a total sample of 44 participants, equally distributed between pathological gamblers and healthy controls, would allow the detection of effect sizes of approximately Coheńs d= 0,76. Importantly, this detectable effect size falls below those reported in previous meta-analyses on emotional reactivity in pathological gamblers assessed with fMRI (^41^), suggesting that the present study is sufficiently powered to detect meaningful between-group differences. Nevertheless, it should be noted that direct comparisons between fMRI and fNIRS findings are not straightforward.

### Participants

Target participants will be adult individuals (over 18 years). The study uses a single-factor design with two levels of gambling experience: 1) a group consisting of individuals with no or low gambling experience (G1), and 2) a clinical group composed of individuals with a history of high-frequency gambling behaviour or gambling disorder (G2).

Participants must meet the following criteria: a) age over 18 years, b) fluency in Spanish, c) provision of written informed consent, d) absence of neurological condition that may affect sensory or cognitive processing (e.g., acquired brain injury, neurodegenerative disorders), e) absence of an active severe psychiatric disorder or developmental condition that may interfere with task performance or data interpretation, f) absence of contraindications for neuroimaging procedures (e.g., metalling implants, cardiac pacemaker. Participants will be excluded if they meet any of the following conditions: a) exhibit excessive movement during fNIRS acquisition, compromising data quality, b) show emotional ratings during the Self-Assessment Manikin (SAM) calibration phase that deviate significantly from the normative valence ratings established in the International Affective Picture System (IAPS), according to predefined deviation thresholds, c) voluntarily show revocation to participate.

### Measures

#### Emotional stimuli

Emotional stimuli consist of images selected from the International Affective Picture System (IAPS), comprising scenes depicting interpersonal interactions with visible faces in order to enhance emotional engagement. Stimuli are selected based on normative valence and arousal ratings, adapted to the Spanish population (^42^). Three categories of images are included, selected from the standardized database of the IAPS: 1) neutral (medium valence, low arousal), 2) sexual (heterosexual nude couples) and 3) violence. Sexual and violence images are expected to elicit comparable levels of arousal while differing in valence (positive and negative, respectively). Neutral images will serve as an emotional baseline condition, with medium valence and low arousal. An additional fourth category, gambling, was created specifically for this study. These images depict gambling-related advertisements and were collected from publicly accessible online sources advertising gambling activities. Gambling stimuli were selected to match IAPS images in terms of visual properties (resolution, luminance, and framing) to reduce perceptual confounds.

#### Physiological measures

Cerebral hemodynamic and metabolic activity will be assessed using a recently developed functional near-infrared spectroscopy (fNIRS) system (Theia model, Newmanbrain S.L., Alicante, Spain), featuring 16 short channels (14mm) and 12 long channels (32mm) (Fig.2). A detailed description of this device can be found in a prior publication from our research group (^43^). Based on the international 10-5 system, the NIRS probe will be positioned on the forehead, aligned with AFpz (Fig.2), primarily covering the rostromedial prefrontal cortex (RMPFC), a region frequently examined in emotion-related research (^44^). The device will be positioned using a flexible headband and adjusted to ensure adequate contact between the sensors and the skin, thereby optimizing signal quality. Due to the pressure required to maintain proper optical coupling, some participants may experience mild discomfort during the measurement.

#### Psychological and behavioral measures

Gambling behavior and related psychological constructs will be assessed using a combination of ad hoc questionnaires and standardized instruments.

– *Gambling frequency*, measured with an ad hoc 8-item questionnaire assessing the number of occasions of gambling over the past 30 days across different modalities, including online sports betting, land-based sports betting, slot machines, online poker, in-person poker with friends, online casino games, roulette, and scratch cards.
– *Gambling intensity,* measured using an ad hoc 8-item questionnaire assessing the amount of money (in euros) spent during the previous 30 days for each gambling modality described above.
– *Gambling severity,* evaluated using the SOGS (South Oaks Gambling Scale) (^45^) Spanish adaptation by Echeburúa (^46^). This scale consists of 15 yes-no items. The original scale demonstrated a high reliability (Cronbach’s α = .97).
– *Impulsivity*, assessed using the Plutchik Impulsivity Scale (^47^) Spanish version validated by Rubio et al. (^48^). This scale consists of 15 items reflecting tendencies toward unplanned or impulsive behaviors, rated on a 4-point frequency scale ranging from “never” to “almost always”. Total scores range from 0 to 45, with scores >20 indicating high impulsivity. The original scale demonstrated acceptable internal consistency (Cronbach’s α = .73).
– *Sensation seeking*, measured using the BSSS-8 short version by Hoyle et al. (^49^). This 8-item instrument assesses four dimensions of the trait (thrill and adventure seeking, experience seeking, disinhibition, and boredom susceptibility), each represented by two items. Responses are provided on a 5-point Likert scale (1=strongly disagree to 5=strongly agree), yielding total scores between 8 and 40. The original scale demonstrated acceptable internal consistency (Cronbach’s α = .75)
– *Alexithymia*, assessed using the TAS-20 (^50^). This self-report instrument assesses three dimensions of the construct: difficulty identifying feelings, difficulty describing feelings, and externally oriented thinking. Responses are provided on a 5-point Likert scale (1=strongly disagree to 5=strongly agree), yielding total scores between 20 and 100, with higher scores indicating greater levels of alexithymia. Internal consistency α = .78
– *Emotional response*, evaluated using the Self-Asssessment Manikin (SAM) (^51^), a visual analog scale consisting of pictographic representations of human figures with graded intensity. This instrument usually assesses two affective dimensions: valence and arousal. Participants rate each image by selecting one of five figures or intermediate positions, resulting in scores ranging from 1 (lowest valence/arousal) to 9 (highest).

### Procedure

#### a Timeline of the study protocol

Participant recruitment will be carried out through community-based recruitment strategies targeting the general population, as well as in collaboration with several addiction treatment associations located in the province of Alicante. These centers provide treatment for substance-related and behavioral addictions, including gambling disorder.

The recruitment phase will take place over a period of two months (Fig.1). During this period, the research team will establish contact with the directors of addiction treatment associations and community organizations in order to present the study objectives and procedures. Upon obtaining institutional approval, potential participants will be informed about the study by association staff following a brief standardized presentation. In parallel, recruitment of participants with no- or low-frequency of gambling will be carried out through community outreach strategies, including dissemination of the study information via institutional social media platforms, the display of printed posters at collaborating centers, and community networks within the same geographical area. Age and sex will be assessed for each potential participant.

**Fig 1.**
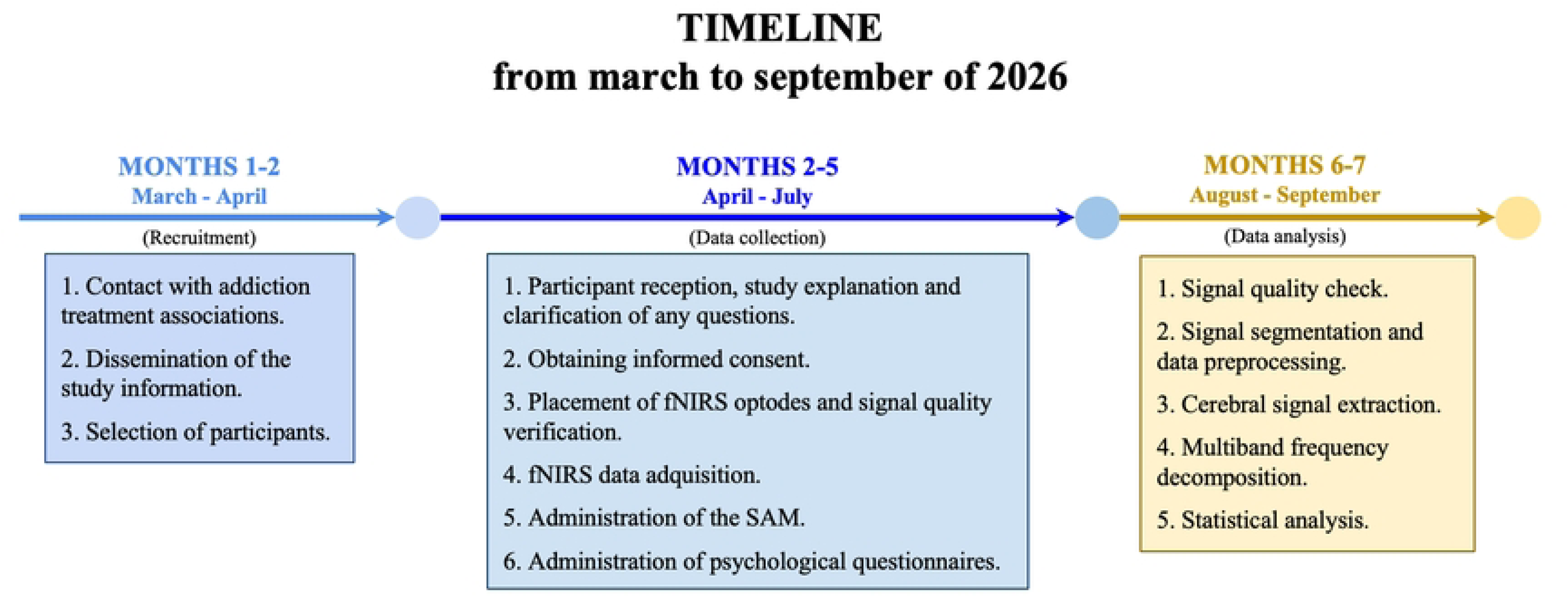
Overview of the timeline of the study protocol

Given that the number of individuals presenting specifically with gambling-related problems varies across centers, a multisite recruitment strategy has been adopted to ensure adequate sample size and representativeness of the clinical population. Control participants will be selected using a purposive sampling strategy and individually matched to clinical participants based on sex and age. Age matching will be conducted exactly when possible or within a ± 2-year range when necessary. Additionally, control participants will be screened to ensure the absence of current or past gambling-related problems or other addiction disorders.

Then, the experimental procedures and data collection will be carried out between months 2 and 5, including participant onboarding, fNIRS data acquisition, and administration of behavioral and psychological measures (Fig.1). Data processing and analysis will subsequently be conducted during months 6 and 7 (Fig.1).

#### b Experimental procedure

Experimental sessions involving fNIRS acquisition will be conducted in a controlled laboratory environment at the Spanish public university Miguel Hernández University of Elche. Participants will be seated comfortably in a softly illuminated, and sound-attenuated room, ensuring standardized lighting, noise and seating conditions to optimize signal quality and minimize artifacts. A single trained experimenter (male psychologist) will supervise all sessions to ensure procedural consistency.

After fNIRS optode placement and signal quality verification, the recording will begin with a 180-second resting baseline during which participants will view a blank screen while remaining relaxed. Following the baseline, seven image blocks will be presented in the following fixed order to all the participants: neutral, gambling, neutral, violence, neutral, sexual and neutral. This fixed order is a deliberate methodological decision aimed at maximizing the internal validity of the design. The literature shows that highly arousing stimuli, such as those with violent or sexual content, can generate persistent physiological and neural responses that can contaminate subsequent conditions (^52^). The fixed order prevents these highly arousing conditions from interfering with the response to the gambling advertising stimuli. In addition, neutral blocks are interspersed between emotionally salient conditions to facilitate emotional recovery and reduce carry-over effects. Each block contains 13 images. Each image will be displayed for 5 seconds, followed by a 1-second blank screen to minimize visual persistence effects. Therefore, each block will last (5 + 1) * 13 = 78 seconds (Fig.2). During image presentation, participants will be asked to perform a simple valence categorization task for each image using two buttons on a response pad, the right for pleasant, the left for unpleasant and neither for neutral/indifferent. This task is intended to ensure sustained attention and active emotional appraisal rather than passive viewing. After completion of the seven blocks, a 180-second recovery period will be recorded under resting conditions.

**Fig 2.**
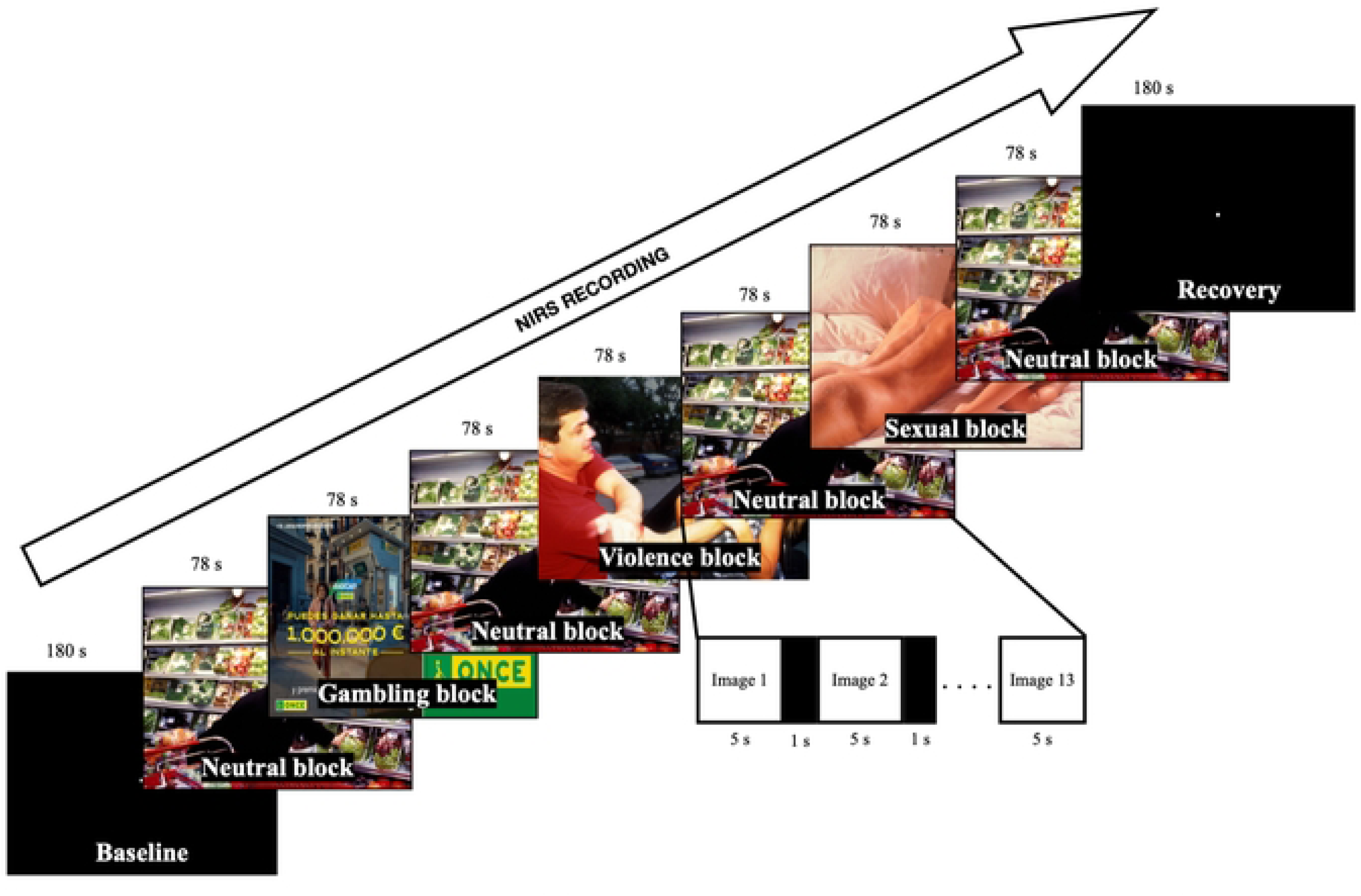
Overview of the experimental design, including the sequence of stimulus blocks and representative images for each condition.

#### c Post-recording behavioral and psychological assessment

After fNIRS data acquisition, participants will be instructed to rate their subjective emotional responses to the presented images using the nine-point SAM scale, to assess both valence and arousal. These ratings will be used to verify emotional engagement and to perform calibration analyses relative to normative IAPS values.

In addition, participants will complete the set of questionnaires relative to psychological measures described above.

### fNIRS data management plan

#### a fNIRS instrumentation and signal quality check

Theia, a NIRS system developed by Newmanbrain S.L. (Elche, Spain), will be employed, featuring 16 short channels (14 mm) and 12 long channels (32 mm) (Fig.3). A detailed description of this device can be found in a prior publication from our research group (^43^). Based on the international 10-5 system, the NIRS probe will be positioned on the forehead, aligned with AFpz (Fig.3), primarily covering the RMPFC, a region frequently examined in emotion-related research (^44^).

**Fig 3.**
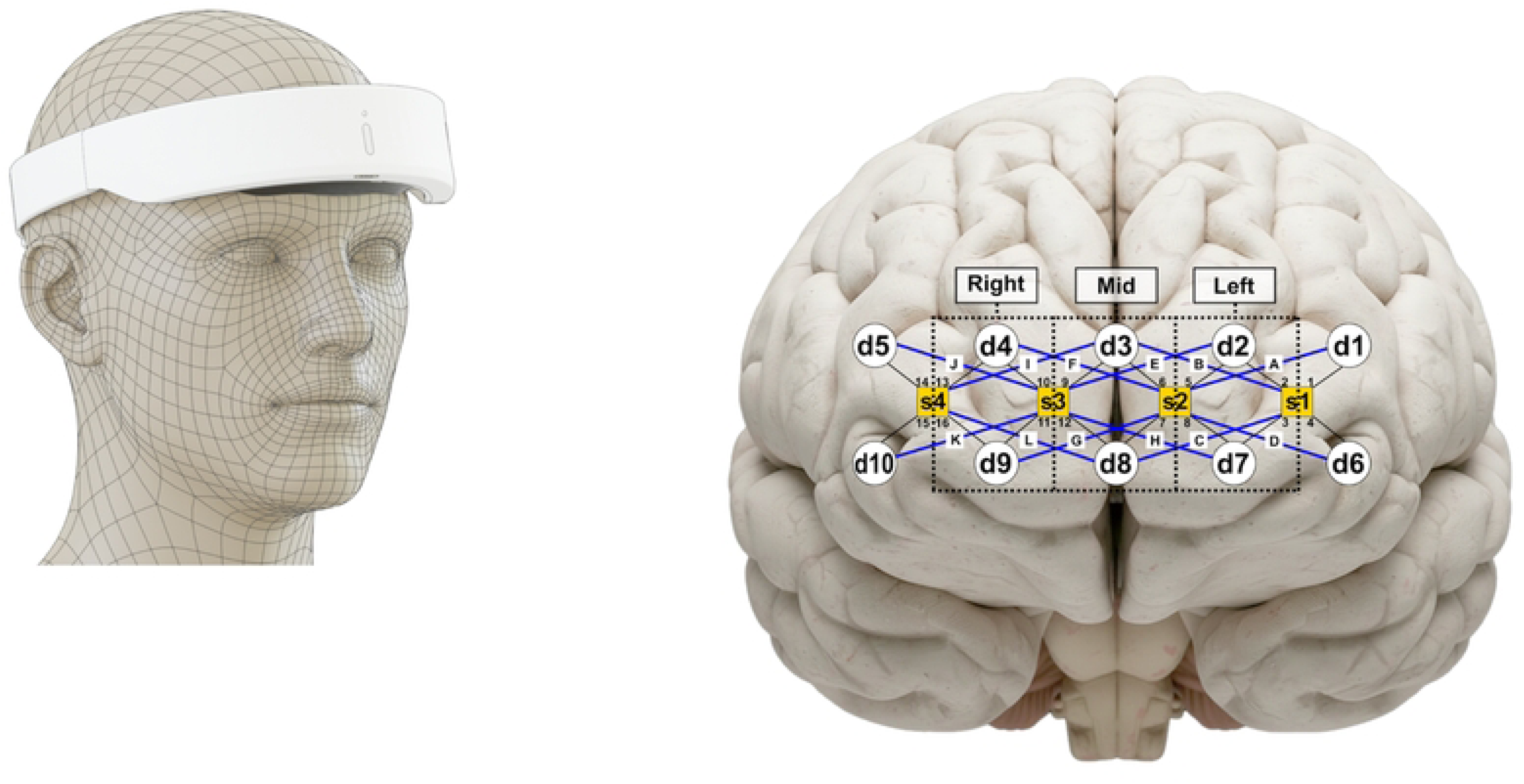
Probe geometry and placement (yellow squares = emitters; white circles = detectors), providing 16 short-channels (black lines with numbers) and 12 long-channels (blue lines with letters).

In order to detect values falling outside the device’s dynamic range or showing excessive coefficient of variation (> 7.5%) (^53, 54^) and exclude low-quality recordings, we will examine the raw optical data. Additionally, recordings affected by motion artifacts will be discarded by identifying abrupt signal changes that coincide with accelerometer spikes.

#### b Signal segmentation and data preprocessing

From the raw optical data, we will extract the 546-s task interval corresponding to the seven picture blocks (78-s each). To reduce boundary effects associated with subsequent processing techniques (e.g. wavelet decomposition) (^55^) an additional 180-s of baseline and recovery data will be included, resulting in a final 906-s data segment (Fig.4). Thus, for each wavelength, 16 raw data segments will be obtained from the short-channels and 12 from the long-channels. All post-processing procedures will be conducted using these data segments, and analyses will be performed in MATLAB (Version R2021b, MathWorks, Natick, MA, USA), employing built-in functions, custom-developed scripts, and open-source toolboxes.

**Fig 4.**
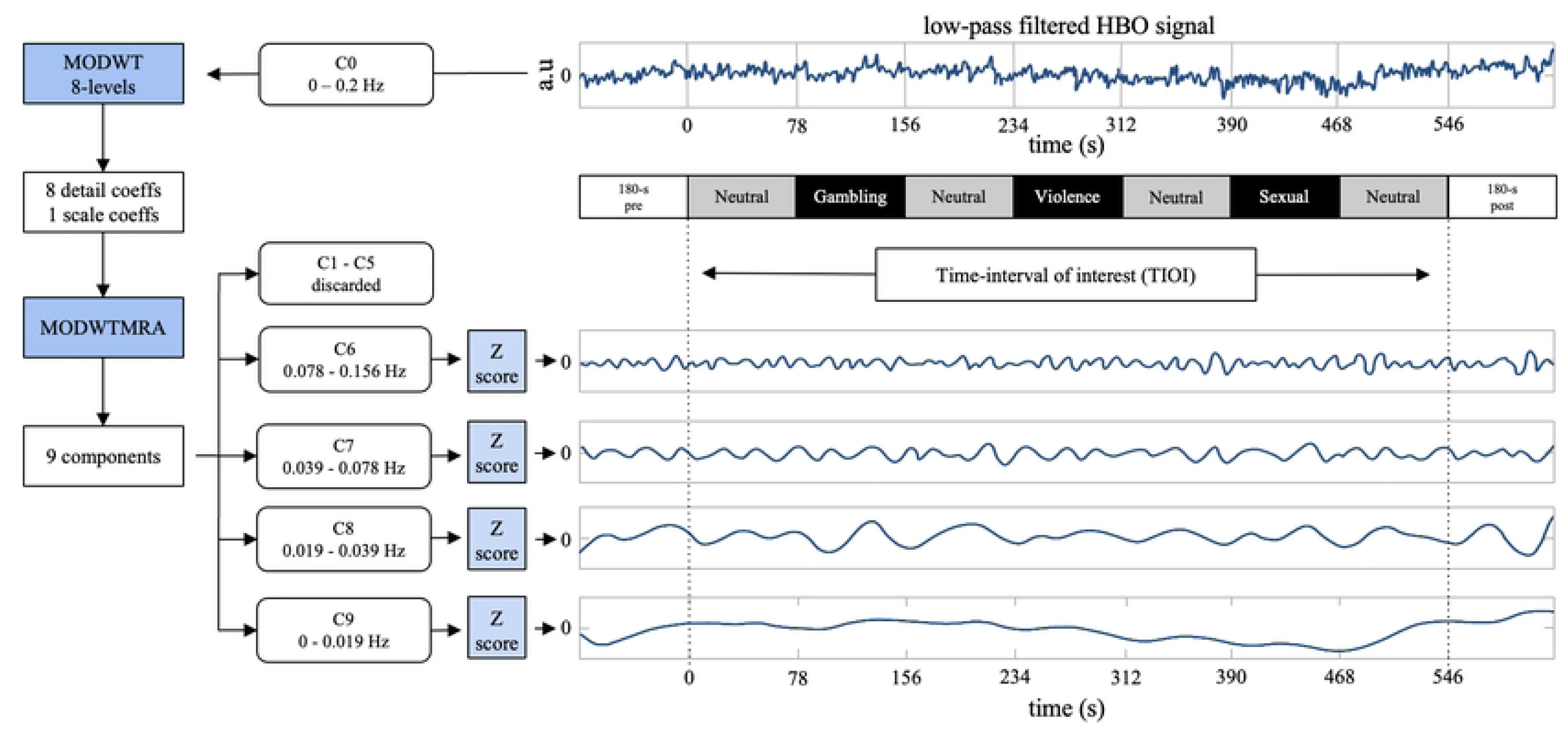
Overview of the frequency decomposition procedure

Relative changes in oxyhemoglobin (HbO) and deoxyhemoglobin (HbR) concentrations will be estimated for each channel using functions from the Homer2 package (^56^), based on the modified Beer-Lambert law (^57, 58^) and applying a differential pathlength factor computed according to Scholkmann and Wolf (^59^). The HbO and HbR signals will subsequently be low-pass filtered at 0.2 Hz using a zero-phase 5th-order Butterworth filter to attenuate respiratory, cardiac, and high-frequency instrumental noise (^56^). Consequently, for each chromophore, 16 time series will be obtained from short channels and 12 from long channels, hereafter referred to as short signals (SS) and long signals (LS), respectively.

#### c Cerebral signal extraction

Deep fNIRS measurements are affected by hemodynamic fluctuations that do not originate in the cerebral cortex (^60, 61, 62^). A widely adopted approach to address this issue is the use of multi-distance measurements (^63, 64^), based on the assumption that short channels primarily capture extracerebral contributions, whereas long channels contain both extracerebral and cortical components (^65, 66^). Accordingly, SS can be used as reference measurements to regress out superficial interference from LS (^67, 68, 69^).

Among the available approaches, regression techniques can be employed under the assumption that physiological noise exhibits correlated temporal dynamics in both SS and LS, whereas task-evoked cortical responses remain independent (^69, 70^). Previous studies have demonstrated that superficial hemodynamics is spatially heterogeneous (^67, 60^), highlighting the importance of selecting reference signals (RS) in close proximity to the long channels to be corrected (^65, 71^). Given that the NIRS system provides up to three SS candidates satisfying the proximity criteria for each LS, a double SS regressor will be implemented by combining two SS into a single RS, following the approach proposed by Gagnon et al. (^72^) and the procedure previously described by our group (^43^). This strategy yielded 12 reference signals, which were used for the regression of the corresponding 12 LS. Following regression, 12 denoised signals will be obtained for both HbO and HbR, hereafter referred to as clean (or corrected) signals (CS), which are assumed to predominantly reflect cortical hemodynamic activity.

Given that the optode placement may vary slightly across participants due to differences in their head anatomy (^73^), strategies to enhance signal robustness are required. In this study, we address this issue by applying a spatial clustering approach, which has been shown to improve signal-to-noise ratio and reliability (^74, 75^). Specifically, to avoid interpreting single-channel fluctuations, signals will be aggregated within three predefined regions of interest (ROIs): left, medial, and right (Fig.3). For each participant, the four leftmost long-channels (A, B, C, D) will be grouped to obtain averaged RS, LS, and CS signals. The same procedure will be independently applied to the medial (E, F, G, H) and rightmost (I, J, K, L) channel groups (Fig.3). As a result, each ROI will be represented by three mean signals corresponding to RS, LS, and CS. Subsequent analyses will be performed on these aggregated signals, which are expected to exhibit comparable signal-to-noise characteristics across ROIs, as each is derived from an equivalent number of neighbouring signals and computed following the regression model: CS = LS − β0 + β1RS. For each ROI, mean time courses of HbO and HbR will be calculated across participants for each signal type, along with the corresponding standard error of the mean (SEM).

#### d Multiband frequency decomposition

Physiological signals are typically nonstationary, meaning that their spectral content may vary over time. In this context, wavelet-based approaches are particularly well suited, as they are sensitive to time-varying dynamics and capable of revealing underlying patterns that may not be apparent with conventional methods. In this protocol, the undecimated maximal overlap discrete transform (WODWT) was selected (^76, 77^), given its suitability for synchronization analysis (^78, 79^) and its widespread use in neuroimaging applications for frequency decomposition (^80^). Following the procedure described previously by our research team in Molina-Rodríguez et al. (^81^), an 8-level decomposition will be performed using the “modwt” function in MATLAB, which implements a Fourier-based version of the MODWT, together with Daubechis wavelet of order 5 (db5) (^82, 83^). This procedure will yield eight sets of detail coefficients along with a final set of scaling coefficients (Fig.4). Subsequently, multiresolution analysis will be carried out using the “modwtmra” function, resulting in nine oscillatory components in the time domain (denoted as C1-C9), whose sum fully reconstructs the original signal. As the “modwtmra” function operates as a zero-phase filter, the extracted components will remain temporally aligned with the original signal, preserving their features on the same time scale.

Given that the upper frequency limit of the original signals will be set at 0.2 Hz, the analysis will focus on components C6 (0.078–0.156 Hz), C7 (0.039–0.078 Hz), C8 (0.019–0.039 Hz), and C9 (0–0.019 Hz). For consistency, the original signal will be denoted as C0, representing the conventional preprocessed signal commonly used in fNIRS studies. Finally, to facilitate comparisons and averaging procedures, all components will be standardized to z-scores when required (Fig.4).

## Statistical Analysis

In the present study protocol, analysis will focus on HbO signals, given their higher sensitivity to superficial hemodynamics in the forehead and their reported correspondence with fMRI BOLD responses in the middle frontal cortex across a range of emotional tasks (^84, 85^).

Following a pairwise framework (^28^), symmetric matrices of Pearson correlation coefficients will be computed for HbO signals across all possible participant pairs, within each of the seven task blocks, and separately for each channel, component (C0, C6-C9), and signal type (RS, LS, CS). ISC will be estimated as the median of the upper triangular elements of each matrix, providing a robust measure of central tendency that avoids the need for Fisher transformation and is suitable for both parametric and nonparametric statistical testing (^86^). Given a total sample size of 40 participants (20 per group), each median will be derived from (40² – 40)/2 = 780 pairwise values, providing a large number of observations for robust ISC estimation (^87^).

To assess statistical significance, a nonparametric two-sample test will be performed using a bootstrapping procedure (^86^). For each iteration, the median ISC will be computed after replacing the rows and columns of the correlation matrix with those corresponding to N participants randomly sampled with replacement, and subsequently centered by subtracting the observed ISC value (^28^). Approximate null distributions will be generated using 10,000 resampling iterations in order to estimate p-values. These p-values will be corrected for multiple comparisons by controlling the false discovery rate (FDR) at q = 0.05 (7 task blocks x 3 ROI = 21 comparisons). In addition, 95% confidence intervals (CI) will be estimated by bootstrapping the upper-triangular elements of the observed correlation matrix (2,000 resamples).

Achieving statistical significance in ISC for a given oscillatory component may be misleading, as it could arise from spurious correlations, particularly at lower frequencies. Therefore, it will be necessary to determine whether ISC values are significantly greater than those observed in other components. To this end, the method described by Kauppi et al. (^79, 88^) will be applied, based on a modified Pearson-Filon statistic using Fisher’s z-transformation (ZPF) (^89^), which is appropriate for testing differences between two non-overlapping dependent correlations (^90^). Briefly, for each pair of components (a, b), ZPF statistics will be computed for all participant pairs within each channel. These values will then be aggregated to obtain summed statistic (ZPFᵃᵇ∑) across all pairs. An approximate permutation distribution of ZPFᵃᵇ∑ will be generated by randomly flipping the sign of individual ZPF values, under the null hypothesis of zero-mean distribution. For each permutation, maximal and minimal statistics across all channels will be retained to construct a symmetric null distribution. This null distribution will be estimated using 20,000 permutations, and significance thresholds will be determined at α = 0.05 based on the extreme values of the distribution, thereby implicitly controlling the family-wise error rate (FWER) for multiple comparisons (^91^). To characterize the response patterns, the average time course of HbO and HbR will be computed for each channel, component, and signal type. Although ISC will not be evaluated for HbR, examining both chromophores is recommended to provide a more comprehensive interpretation of fNIRS responses (^92^).

To examine the relationship between psychological variables associated with pathological gambling, multiple regression analyses will be conducted in which ISC values under the gambling condition will be entered as the dependent variable, and scores from the psychological assessment measures will be included as predictors. In addition, to explore potential differences in emotional responses, repeated-measures ANOVAs will be conducted on SAM rating, analysing valence and arousal across pictures categories, with “picture category” included as a within-subject factor.

## DISCUSSION

The present study protocol is designed as a cross-sectional comparative investigation with the objective of exploring the neural and behavioral impact of gambling-related advertising using fNIRS combined with ISC analysis. Specifically, the study aims to validate the hypothesis that populations with a high-frequency of gambling behaviors process gambling cues in a more idiosyncratic and heterogeneous manner (H1 and H4), while maintaining normative responses to other high-arousal stimuli, such as violent or sexual content (H2). Ultimately, this approach seeks to assess the predictive capacity of psychological variables on neural engagement (H3), providing a multidimensional exploratory framework of gambling behavior.

To date, neuroimaging research on gambling advertising has predominantly relied on fMRI, which often lacks the ecological validity and temporal resolution needed to capture rapid affective processing in naturalistic settings (^18, 19, 20^). Therefore, our project, by employing a portable fNIRS system and a data-driven ISC approach, represents a pioneering effort to identify objective neural markers of vulnerability to gambling cues. The findings could provide valuable insights for developing public health policies and more effective advertising regulations specifically tailored to protect vulnerable populations.

Regarding the study management, any necessary amendments to the protocol (e.g., changes in recruitment strategies or technical adjustments) will be formally documented and submitted to the Ethics Committee for Research Involving Medicines at the University Hospital of San Juan - Alicante and the Research Ethics Integrity Committee of Miguel Hernández University for approval prior to implementation. In the unlikely event of study termination due to insufficient recruitment, safety concerns, or unforeseen technical failures, all data collected up to that point will be securely archived, and the reasons for termination will be transparently reported in any subsequent publications. In this concern, it is worth to note that at the time of this protocol’s publication, the recruitment phase has already commenced and preliminary tracking indicates that the flow of participants and the available sample pool from the collaborating centers are sufficient to meet the target sample size requirements.

Nevertheless, our study presents several difficulties and strengths that must be acknowledged. First, the success of this study relies on the proper recruitment of a clinical or near-clinical population. Given the challenges in reaching individuals with high-frequency gambling behavior or gambling disorder, we have outlined several measures to ensure an adequate sample size. Specifically, 1) we have adopted a multisite recruitment strategy in collaboration with specialized addiction treatment centers; 2) G1 participants will be individually matched to G2 participants based on sex and age to minimize demographic confounding; and 3) we have conducted a sensitivity power analysis to ensure the study is sufficiently powered to detect meaningful between-group differences.

Despite these efforts, the generalizability of our findings may be limited. First, preliminary tracking of the ongoing recruitment indicates a significant gender imbalance, with the sample pool consisting exclusively of men. While this reflects the epidemiological reality that the prevalence of gambling disorder and high-frequency gambling is substantially lower among women (^93, 14^), it restricts the generalizability of our results to the female population. Second, the use of static images as emotional stimuli may not fully capture the complexity of modern gambling advertising, which typically employs multi-sensory audiovisual narratives; future research should consider incorporating auditory components to enhance ecological validity. Third, although the fixed sequence was a deliberate choice, potential presentation order effects may not be fully controlled, so exploring alternative presentation orders or counterbalancing techniques in future investigations would enhance the generalizability and robustness of the further observed effects. Finally, as previously noted, the fNIRS probe is primarily restricted to the RMPFC, meaning that deeper subcortical structures involved in reward processing (e.g. the striatum) cannot be directly monitored. Integrating fNIRS with subcortical recording techniques could provide a more holistic view of the gambling brain, though researchers must carefully balance the gain in spatial depth with the constraints of a less naturalistic environment (19, 20, 21).

A primary strength of this protocol is the use of standardized, validated assessment tools and advanced signal processing techniques (^37, 43^). In particular, the implementation of short-channel regression to minimize extracerebral noise and the use of the IAPS as a benchmark for emotional engagement will enhance the reliability of our measurements. Although psychological measures are self-reported and may be subject to social desirability or recall biases, the inclusion of neurobiological data through fNIRS allows for a more robust and objective evaluation of the emotional impact of advertisements.

In conclusion, this protocol establishes a methodological framework to better understand the neural responses to gambling-related advertising. All findings generated from this research will be disseminated through peer-reviewed publications, presentations at international conferences, and reports to clinical and educational associations, by which we aim to bridge the gap between neuroscientific research and its practical application. Ultimately, this project seeks to provide a perspective on how marketing influences addictive behaviors, potentially supporting the future design of evidence-based preventive measures and public health strategies.

## Contribution (CRedIT Taxonomy)

Conceptualization: JIB, DLLI, Data curation: DLLI, Formal analysis: JIB, SMR, Funding acquisition: DLLI, Investigation: ARL, SMR, Methodology: JIB, DLLI, SMR, Project administration: DLLI, Resources: JIB, OPP, Software: JIB, Supervision: DLLI, Validation: SMR, Visualization: ARL, SMR, Writing – original draft: ARL, SMR, DLLI, Writing – review & editing: VCP

## Data Availability

No datasets were generated or analysed during the current study. All relevant data from this study will be made available upon study completion.

## Acknowledgement

The authors would like to thank the associations GAEX (Torrevieja), Nueva Vida (Torrevieja and Villena), and APAEX (Elche) for their collaboration and support. The authors also thank Miguel Ibáñez Ballesteros for his assistance with the preparation of images and figures.

